# Equity in Paediatric Emergency Departments during COVID-19

**DOI:** 10.1101/2020.09.25.20201533

**Authors:** Sophie Thorne, Sunil Bhopal, Christian Harkensee, Alex Battersby, Anna Brough, Stephen Owens

## Abstract

Children’s attendances in paediatric emergency departments have fallen precipitously in North East England and elsewhere in recent months. We analysed data from 3 hospitals to understand which children were not being brought during the COVID-19 ‘lockdown’. In our population there is no evidence of a disproportionate impact on children belonging to vulnerable sociodemographic groups and no obvious change in illness acuity among those children still attending. However we noted a marked reduction in infectious disease presentations which might reflect one positive impact of enhanced social distancing on child health. More granular data describing the ‘collateral damage’ of the COVID-19 pandemic to children’s clinical services are needed to plan for the mitigation of its continuing effects.

**What is known on this topic:**

1. Presentations to paediatric emergency departments in Europe and the United States have reduced dramatically during the COVID-19 pandemic ‘lockdown’.

**What this paper adds box:**

1. This is the first paper to show that reduced attendance was proportionate across different deprivation and ethnicity groups.
2. We show that presentations of children with infectious diseases reduced more than those with other conditions or trauma.
3. There was no change in admission rates, taken as a broad indicator of illness acuity at presentation among the population still attending paediatric emergency departments.

## Introduction

By 1 March 2020 SARS-CoV-2 was spreading throughout the United Kingdom (UK). In response, the UK government announced a series of progressive social distancing measures culminating in full ‘lockdown’ from 23 March 2020, with instructions to ‘Stay at Home, Protect the NHS, Save Lives’. Whilst travel to receive medical attention was permissible under these regulations, clinicians in Paediatric Emergency Departments (PED) became increasingly concerned about falling attendances and the danger that sick and injured children might not be brought for urgent healthcare. In late-March 2020 there was more than a 60% reduction in the number of PED attendances in Greater Manchester compared to the same period in 2019 [2]. In Italy, which implemented lockdown earlier than the UK, a comparable decrease of 73-88% was associated with reports of deaths in children following delayed presentations of sepsis and diabetic ketoacidosis [3].

In light of these reports and in the absence of greater granularity to the data, we were particularly concerned that children from disadvantaged sociodemographic groups would be accessing healthcare even less commonly than those from more privileged backgrounds, and that presentations of specific conditions, especially infections, might be delayed. Evidence for such a change would provide the basis for targeted local and national health-messaging, aimed at groups at greatest risk.

## Methods

We extracted and compared routinely-collected hospital episode data on each child (0-18 years) who attended PED at Great North Children’s Hospital (CH) between January and April 2019 and the same period in 2020. The Great North Children’s Hospital is the tertiary referral centre for children in the North East of England and North Cumbria and one of 14 children’s hospitals in the UK. In addition we extracted the same data for the two nearest district general hospitals (DGH) with PED which refer children to GNCH: Queen Elizabeth Hospital, Gateshead (DGH1) and Northumbria Specialist Emergency Care Hospital, Northumberland (DGH2). Together these three units serve an estimated total population of almost 200000 children. Caldicott approval for the data extraction was obtained from each site. Formal ethics approval was not required per NHS Health Research Authority decision-tool (http://hra-decisiontools.org.uk/research/).

We calculated the number of children attending each PED per calendar month. We compared attendance by age, gender, ethnicity, index of multiple deprivation decile (IMD) and the income deprivation affecting children index (IDACI) score, calculated for each child on the basis of postcode [4]. We recoded captured ethnicity data into the six groups recognised by the UK Office for National Statistics [5]. To understand common presentations, ST and SO grouped the SNOMED CT diagnostic descriptors of presenting children *pre-hoc* into six broad categories: Surgical (Trauma and Non-trauma), Medical (infectious and non-infectious), Psychiatric and Non-discriminatory. We used admission rate as a crude marker of illness acuity in presenting children. We analysed data for each unit individually and then all three units combined and present descriptive statistics. Null hypothesis testing for the changes in proportion of attendances was done using chi-squared tests and 95% confidence intervals (CI) are presented around the estimated effect sizes.

## Results

There were 16154 attendances from 1 January 2019 to 30 April 2019 across all three units (3456 CH; 6921 DGH1; 5777 DGH2). Attendances fell to 11975 in the same period in 2020 (2625; 5442; 3908 respectively). The number of attendances in April 2020 was 63% (95% CI 60, 65.6) lower than in April 2020. This pattern was similar across all three units (Figure 1).

**Figure 1:**
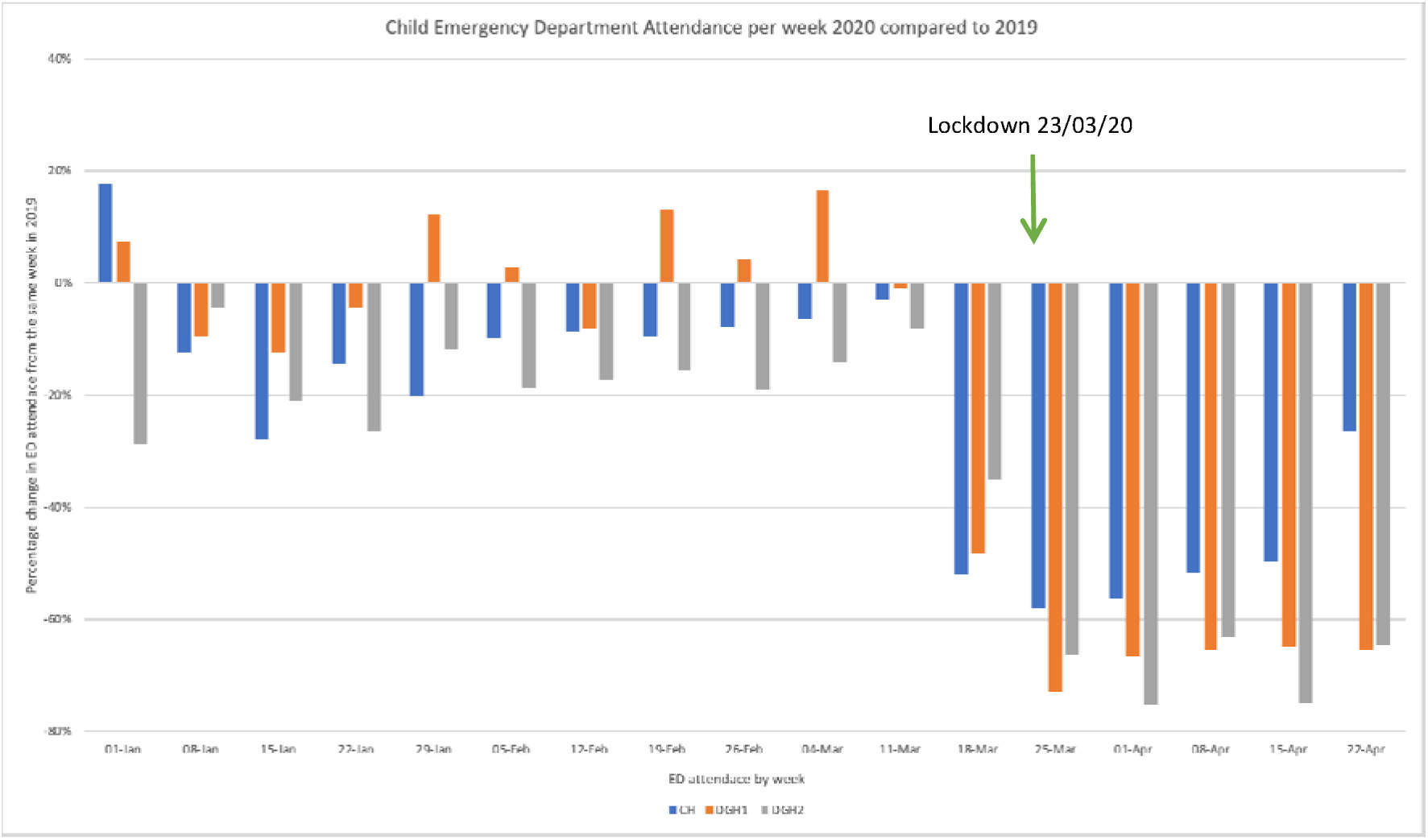
Change in Child Emergency Department Attendance 2019 to 2020.

There were no differences in the proportions of children attending from the highest deprivation deciles, Black, Asian and Minority Ethnic (BAME) groups or by gender between April 2019 and April 2020 (Table 1). We noted a small increase in the proportion of pre-school aged children presenting (0-5 years). There were no differences in any of the demographic indicators in the under-5 age group (data not shown).

**Table 1:** Summary Table of Demographics of Children Presenting to Emergency Departments in the North East of England.

|  |  |  |  |  |  |  |  |  |
| --- | --- | --- | --- | --- | --- | --- | --- | --- |
| AGE ≤5 | CH | 417 | 54.9% | 232 | 60.6% | 5.7% | [-2.2, 13.6] | 0.07 |
|  | DGH1 | 783 | 47.8% | 278 | 49.6% | 1.9% | [-5.0, 8.7] | 0.445 |
|  | DGH2 | 818 | 61.1% | 303 | 69.3% | 8.2% | [2.0, 14.4] | 0.002 |
|  | ALL | 2018 | 54.0% | 813 | 58.9% | 4.9% | [0.9, 8.9] | 0.002 |
| BAME | CH | 179 | 23.6% | 81 | 21.2% | -2.4% | [-13.2, 8.5] | 0.36 |
|  | DGH1 | 218 | 13.3% | 69 | 12.3% | -1.0% | [-9.9, 8.0] | 0.553 |
|  | DGH2 | 63 | 4.7% | 20 | 4.6% | -0.1% | [-10.7, 10.4] | 0.91 |
|  | ALL | 460 | 12.3% | 170 | 12.3% | 0.0% | [-5.8, 5.8] | 0.993 |
| FEMALE | CH | 317 | 41.7% | 184 | 48.0% | 6.3% | [-2.7, 15.4] | 0.04 |
|  | DGH1 | 781 | 47.7% | 259 | 46.3% | -1.4% | [-8.4, 5.6] | 0.566 |
|  | DGH2 | 597 | 44.6% | 204 | 46.7% | 2.1% | [-5.9, 10.0] | 0.452 |
|  | ALL | 1695 | 45.4% | 647 | 46.9% | 1.5% | [-3.0, 6.0] | 0.331 |
| IMD DECILE 1 AND 2 | CH | 327 | 43.0% | 170 | 44.4% | 1.4% | [-7.8, 10.6] | 0.66 |
|  | DGH1 | 696 | 42.5% | 219 | 39.1% | -3.4% | [-10.8, 4.1] | 0.164 |
|  | DGH2 | 407 | 30.4% | 144 | 33.0% | 2.5% | [-6.4, 11.4] | 0.32 |
|  | ALL | 1430 | 38.3% | 533 | 38.3% | 0.0% | [-4.5, 5.2] | 0.816 |
| IDACI DECILE 1 AND 2 | CH | 338 | 44.5% | 174 | 45.4% | 1.0% | [-8.1, 10.1] | 0.76 |
|  | DGH1 | 662 | 40.5% | 211 | 37.7% | -2.8% | [-10.3, 4.8] | 0.247 |
|  | DGH2 | 432 | 32.3% | 150 | 34.3% | 2.0% | [-6.8, 10.8] | 0.431 |
|  | ALL | 1433 | 38.4% | 535 | 38.8% | 0.4% | [-4.4, 5.3] | 0.783 |

Across all three sites, the proportion of children presenting with infectious diseases in April 2020 was 14.7% compared with 28.4% in April 2019 (difference 13.7% (95% CI [8.1, 19.2%]; p<0.001). In the under-5s, this reduction was much larger (21.3%, 95% CI [14.3, 28.3%]; p<0.001). There were no changes in proportions of children presenting with Medical (non-infectious) or Surgical diagnoses overall, however in the under-5s the proportion of trauma cases increased by 13.2% (95% CI [6.4, 19.9%]; p<0.001).

The proportion of children attending PED and subsequently being admitted to hospital was similar between April 2019 and 2020 (26.1% and 25.5% respectively; difference -1.1%; 95% CI [-6.4, 4.3%]), despite the overall fall in attendance s. Almost all admissions were for short-stay assessment only. Results were similar for under-5s and for those attending with infectious illnesses only.

## Discussion

Paediatric emergency department attendance dropped sharply across the three PED examined in the North East of England during the initial stages of the COVID-19 pandemic. During April 2020 the impact was uniform across deprivation deciles, ethnicity and gender but there was a slight over-representation of younger children. Infectious disease presentations fell disproportionately while overall admission rates were unchanged.

Reduced PED attendance during lockdown has been widely reported [2,3]. Our analysis is the first to demonstrate that this effect was proportionate across broad sociodemographic groups. The direct impact of the COVID-19 pandemic has otherwise affected BAME and poorer populations more severely, requiring a more a targeted response in these groups.

Less frequent interactions between children and improved hygiene practices limit transmission of viruses aside from SARS-CoV-2. Fewer childhood infections during lockdown may have accounted for some reduction in PED attendances. Additionally, adherence to ‘Protect the NHS’ messaging augmented by integrated ‘NHS 111’ telephone advice may have led carers to manage most episodes of childhood illness at home. There are concerns that such behaviour might result in children being brought late and more severely unwell to PED [3]. However the fact that the subsequent admission rate was unchanged suggests that this was not a systemic problem in our population.

Study strengths include the large sample size and sampling across three hospitals. The level of deprivation was high, giving a clear opportunity to examine this concern. However the proportion of children from minority ethnic groups was low and this analysis should be repeated elsewhere. A further limitation to interpretation is that around 1.5% of children from both DGHs were later referred or presented to the GNCH. Information-governance safeguards limited our ability to identify and remove these children from dual-analysis.

Clinicians and policymakers might be reassured that whilst admissions are falling, there is no evidence at this early stage, in the North East of England, that particular groups of children have been specifically disadvantaged. There is early evidence that young children were brought for care at an appropriate point in their illness. However future studies might examine specific illnesses in more detail – for example, first presentations of diabetes mellitus type 1 where avoidance of delay is crucial.

Given that social distancing measures are likely to continue for some time, our findings provide initial evidence for planning of emergency healthcare services to help mitigate the ‘collateral damage’ that the pandemic is causing to children. Further work to understand how parents rationalise healthcare seeking behaviour for their children during the pandemic is ongoing in our region (tinyurl.com/CoVID-northeast) and across the UK.

## Data Availability

Available from corresponding author on reasonable request by scientific colleagues

